# Multi-matrix copper exposure is associated with reduced olfactory bulb volume and odor sensitivity in adolescents

**DOI:** 10.64898/2026.07.13.26357935

**Authors:** Michelle A. Rodriguez, Azzurra Invernizzi, Francesca Saviola, Giorgio Pio Marinelli, Kristie Oluyemi, Elza Rechtman, Daniele Corbo, Stefano Renzetti, Cheuk Y. Tang, Lorella Mascaro, Claudia Ambrosi, Roberto Gasparotti, Donald R. Smith, Robert O. Wright, Roberto G. Lucchini, Donatella Placidi, Christoph van Thriel, Megan K. Horton

## Abstract

Copper (Cu) is an essential metal involved in neurobiological processes including energy metabolism and neurotransmission, yet dysregulated Cu levels may adversely affect brain health and olfactory performance. Although olfactory dysfunction has primarily been studied in older adults and neurodegenerative disease, adolescence is a critical period of brain maturation during which the olfactory system may be particularly vulnerable. This cross-sectional study examined associations between Cu exposure, olfactory bulb (OB) volume, and olfactory performance in 200 adolescents and young adults (64% female; ages 13–25) from the Public Health Impact of Metals Exposure cohort. Cu concentrations in blood, urine, hair, and saliva were measured using inductively coupled plasma mass spectrometry. T2-weighted magnetic resonance imaging scans estimated left, right, and total OB volumes using a three-stage deep learning pipeline. Olfactory performance was assessed using the Sniffin’ Sticks test. Weighted quantile sum regression evaluated associations between a Cu mixture index and OB outcomes, while standard linear regression models assessed individual Cu biomarkers. Models were adjusted for age and sex. A higher Cu index was associated with reduced left (β = −0.72, 95% CI [−1.42, −0.02]), right (β = −0.79, 95% CI [−1.43, −0.15]), and total OB volume (β = −1.55, 95% CI [−2.85, −0.25]), as well as lower odor threshold scores (β = −0.23, 95% CI [−0.42, −0.03]). Individual biomarkers were not independently associated with outcomes. These findings suggest that Cu exposure may adversely affect olfactory neurodevelopment during adolescence and highlight the importance of studying environmental exposures relevant to long-term neurological health.

## 1. Introduction

Adolescence is a critical period of continued brain development and maturation marked by refinement of sensory pathways with heightened vulnerability to environmental toxicant exposures. Among brain regions relevant to environmental exposure, the olfactory bulb (OB) is uniquely positioned at the interface of the environment and the central nervous system, serving as the initial site of odor processing and one of the most environmentally exposed neural structures. The development of the OB begins prenatally with neurogenesis and laminar organization of neurons required to transmit sensory information to the brain^1^. Functional odor processing is present at birth indicating that the fundamental structure of the OB is largely established prenatally and maintained postnatally, with minimal evidence for ongoing neurogenesis in humans^2,3^. Despite this early structural establishment, the OB continues to undergo dynamic changes during puberty, with evidence that OB volume and olfactory function increase in association with pubertal timing ^4^. This is likely due to hormonally driven processes rather than the generation of new neurons. The OB is a metabolically active structure due to continuous synaptic activity and sensory processing^5,6^. It receives direct input from the olfactory epithelium and functions as a key neural target for environmental exposures that may enter the brain via the olfactory system^5,7–9^. Studies also show that with increasing age, OB volume and function declines^3,10–13^.

Copper (Cu) is an essential trace metal required for energy production, neurotransmitter synthesis, iron homeostasis, and cellular signaling^14–16^. Dysregulation of Cu homeostasis can induce oxidative stress and neuronal cell death, leading to neurotoxicity including increased risk for neurodegenerative diseases (i.e., Alzheimer’s Disease)^17–19^. Cu readily crosses the blood brain barrier and distributes unevenly across the brain, with concentrations fluctuating across development^20^. Animal studies demonstrate that elevated Cu exposure impairs olfactory function. In coho salmon, Cu exposure disrupts odor-mediated behaviors including predator avoidance, foraging, and social interaction^21,22^. In rodents, Cu dyshomeostasis, led to higher latency periods of food detection through their olfactory sensory function compared to controls^23^. No population-based epidemiologic studies have examined whether Cu exposure influences OB structure or olfactory performance in humans. Emerging clinical evidence from Wilson’s disease, a disorder of Cu accumulation, suggests that disrupted Cu homeostasis may impair olfactory function in humans^24^. However, this relationship has not been examined in large population-based studies particularly during adolescence, a period when the olfactory system undergoes functional refinement and may be especially sensitive to neurotoxicants exposures.

The OB’s anatomical position makes it a biologically plausible target of Cu-related neurotoxicity. Airborne Cu particles can enter the nasal cavity, deposit on the olfactory epithelium, and travel directly to the OB through the olfactory nerve, bypassing the blood brain barrier^8,25^. Magnetic resonance imaging (MRI) studies demonstrate that OB volume is a robust neural correlate of olfactory function, with reduced volume observed in conditions involving olfactory impairment, congenital anosmia, and neurodegenerative disease^26–30^. High-resolution coronal T2-weighted MRI (T2w-MRI) enables precision quantification of OB volume,^31,32^ providing a noninvasive tool for detecting neurotoxicant-associated structural alterations.

Because Cu distributes differently across biological compartments, and each biomarker reflects a distinct physiological process, no single matrix can capture total Cu body burden^33,34^. Blood and urinary markers primarily reflect more recent exposure and excretion, while hair and saliva can integrate longer term metal accumulation^35–37^. As a result, individual biomarkers provide complementary, rather than interchangeable, information about Cu toxicokinetics. Mixtures-based approaches such as weighted quantile sum (WQS) regression offer a principled framework for integrating multiple biomarkers of Cu exposure, each reflecting different aspects of toxicokinetics, into a single exposure index that better represents cumulative internal dose. By leveraging the shared and unique information across multiple matrices, WQS can assist exposure characterization and allows identification of the matrices that contribute most strongly to associations with neurodevelopmental outcomes^33,38,39^.

In this study, we investigate whether Cu exposure is associated with OB structure and olfactory performance in adolescents participating in the Public Health Impact of Metal Exposure (PHIME) cohort. We quantified OB volume using an automated deep learning pipeline applied to high resolution T2-weighted images and assessed olfactory performance using the Sniffin’ Sticks identification test. We measured Cu concentrations in four biological matrices (saliva, blood, hair, and urine) and combined them into an integrated Cu exposure index using WQS regression. We hypothesize that higher Cu exposure will be associated with reduced OB volume and poorer olfactory performance, reflecting early neurostructural and sensory consequences of Cu dysregulation during adolescence.

## 2. Methods and Materials

### 2.1. Participants

The Public Health Impact of Metals Exposure (PHIME) study in Northern Italy investigates associations between metal exposure from anthropogenic emissions and developmental health outcomes in adolescents and young adults. Details of the study have been described elsewhere^40,41^. Inclusion criteria included: birth in the areas of interest; family residence in Brescia for at least two generations; residence in the study areas since birth. The exclusion criteria included: having a neurological, hepatic, metabolic, endocrine or severe psychiatric disorder; using neuroactive medications; having clinically diagnosed motor deficits or cognitive impairment and having visual deficits that are not adequately corrected. Further details on the recruitment process and study design can be found in previous publications^41,42^.

Between 2016-2021, a subset of the PHIME second wave participants (n = 207, ranging from ages 13-25 years old) were enrolled into a follow-up study, PHIME2-MRI, and completed multi-modal magnetic resonance imaging (MRI) scans including a high resolution T2w-MRI scan of the olfactory bulb. PHIME2-MRI participants also provided biological specimens (i.e., saliva, blood, hair, and urine) for metals analysis^38^ and completed an olfactory performance assessment (i.e., Sniffn’ Stick)^41^. All participants met eligibility criteria for MRI scanning including no metal implants or shrapnel, claustrophobia, no prior history of traumatic brain injury, and body mass index (BMI) ≤ 40.

Out of the 207 participants, 200 (97%) participants had usable T2w-MRI images. From the remaining participants, 193/200 (97%) participants had complete exposure data (i.e., Cu concentrations in all 4 matrices; Cu concentrations were incomplete for 7 participants). Following imputation, a total of 200 participants were included in the analysis with complete T2w-MRI scans, behavioral data, and covariates.

Written informed consent was obtained from parents, while participants provided written assent. Study procedures were approved by the Institutional Review Board of the University of California, Santa Cruz and the ethical committees of the University of Brescia, and the Icahn School of Medicine at Mount Sinai.

### 2.2. Biomarker measures of exposure

Venous whole blood, spot urine, saliva and hair biological specimens were collected from each participant, as previously described^40,43–45^. Metal concentrations were measured at the University of California, Santa Cruz, using magnetic sector inductively coupled plasma mass spectroscopy (Thermo Element XR ICP-MS)^40,43–45^. A complete overview of biomarkers can be found in Supplementary Material (Figure S1 and S2). Seven participants were missing one or more Cu biomarkers. We used the chained equations method via the *mice* package in R to handle missing exposure data^46^. This approach estimates missing values by leveraging the relationships among variables in the dataset and accounts for the uncertainty introduced by the imputation process. Geometric mean and standard deviation of Cu in biological samples are presented in Table 1. Participants with missing exposure data did not differ significantly in demographic characteristics relative to those with complete exposure data (n = 193), and no statistically significant differences were observed between the original and imputed Cu distributions (Supplementary Material Table S1).

**Table 1.** Demographics, copper exposure measured through biospecimens, and olfactory performance outcomes measured through Sniffin’ Sticks of adolescents in the PHIME cohort who were included in the current study.

| Characteristics | PHIME participants<br>(n=200) |
| --- | --- |
| **Age (years)** |  |
| mean ± SD | 19.11 ± 2.44 |
| **Sex (n, %)** |  |
| Male | 91 (46%) |
| Female | 109 (64%) |
| **Cu concentrations (GM ± GSD)\*** |  |
| Saliva (ng/mL) | 8.60 ± 2.34 |
| Blood (ng/mL) | 586.34 ± 1.30 |
| Hair (ug/g) | 10.00 ± 1.61 |
| Urine (ug/mL) | 5.92 ± 1.87 |
| **Sniffin' Sticks (mean ± SD)** |  |
| Threshold | 9.46 ± 2.91 |
| Discrimination | 12.77 ± 1.84 |
| Identification | 12.77 ± 1.59 |
| TDI (global olfactory score) | 35.00 ± 3.86 |

**Note:** \*Includes imputed Cu concentrations for all biological matrices. GM = geometric mean, GSD = geometric standard deviation, SD = standard deviation.
*4.2. Cu exposure and olfactory bulb volume*

### 2.3. Olfactory Assessments: Sniffin’ Sticks

To assess olfactory performance, the Sniffin’ Sticks test^47,48^ (Burghardt®, Wedel, Germany) was administered to participants by a trained psychologist. This test consists of three subtests: threshold, discrimination, and identification (TDI)^47^. Generally, the test involves the use of felt pens, the tips of which are saturated with 4 mL of a supra-threshold odorant concentration, dissolved in propylene glycol and antibacterial agent. The odor pen was presented to the participant once, for no more than 3-4 seconds. A single nostril test was performed, holding the pen 2 cm centrally in front of both nostrils. The participant was asked to smell using a simple verbal command (e.g., using the word “attention”) and the interval between each odor pen was around 30 seconds. An interval of 3 to 5 minutes was observed between the three subtests. In the threshold subtest, participants were presented sets of triplet pens containing different dilutions of N-butanol or phenylethyl alcohol according to decreasing concentration; only one pen of the triplet contains the odorant while the other two are solvent-only pens. Participants were tasked with distinguishing between the odorant and solvent pens, with responses recorded as either correctly identified or not identified. A total of 7 points could be obtained. In the Discrimination subtest, participants were blindfolded and presented with 16 sets of three pens: two with the same odor and one with a different odor. Participants had to identify which pen had the different odor. This test was scored based on the correct number of responses out of 16. In the identification subtest, participants are presented with 16 odor pens and were asked to identify the smell of each odor pen. This was administered in a multiple-choice format where they were given 4 answer choices for each pen and asked to select the option that best describes the smell. The identification score represents the number of correct responses. Lastly, a global olfactory score was calculated, consisting of all scores from the 3 subtests (i.e., the TDI score). A TDI score was calculated as the sum of the three subtests and was analyzed as a continuous variable. Normative classifications based on age and sex specific reference values have been described elsewhere^12^.

### 2.4. MRI data acquisition

Magnetic resonance imaging (MRI) data acquisition was performed on a high-resolution 3-Tesla SIEMENS Skyra scanner using a 64-channel head and neck coil at the Neuroimaging Division of ASST Spedali Civili Hospital of Brescia. For each participant, a high-resolution 3D T2w structural MRI scan was acquired (TR =5000ms, TE= 100ms, 72 coronal slices with slice thickness = 2.5mm). Padding was used for the participant’s comfort and to reduce head motion. Earplugs were used for noise reduction. Data were reviewed by a board-certified radiology, performing quality control and checking for incidental findings. No incidental findings were reported.

### 2.5. Olfactory Bulb: Segmentation and Quantification

To localize, segment, and estimate olfactory bulb (OB) volumes, we applied a previously validated deep learning pipeline specifically developed for high-resolution T2w MR images^49^. This fully automated, three-stage pipeline performs end-to-end analysis of 3D volumes in under a minute, making it well-suited for large cohort studies. The pipeline includes (1) OB localization using FastSurferCNN^50^, (2) segmentation within the localized region using an attention-augmented deep learning model (AttFastSurferCNN), and (3) ensemble-based label refinement. This method has already demonstrated high segmentation accuracy, robust generalizability across datasets, and efficient performance, enabling rapid and reliable OB volume quantification at scale^49^. An overview of the volumetric outcomes produced by this pipeline can be found in Supplementary Table S2.

## 3. Statistical Analysis

### 3.1. Descriptive statistics

Visual inspection and descriptive statistics (geometric mean and geometric standard deviation) were used to characterize the metal concentrations in different media. All descriptive statistical analyses were performed using R version 4.5.2.

### 3.2. Individual Cu biomarkers, OB volumes, and olfactory performance

To examine linear associations between Cu concentrations in each individual matrix (i.e., saliva, blood, hair, urine) and olfactory metrics (i.e. OB volumetric and Sniffin’ Stick outcomes), we used standard linear regression models adjusted for age and child sex, adjusting for multiple comparisons (Bonferroni correction). These results can be found in the Supplementary Material (Tables S3 and S4; Figure S4).

### 3.3. Multi-matrix Cu biomarker, OB volumes, and olfactory performance

To estimate associations of multi-matrix Cu exposure and olfactory outcomes (OB volume and Sniffin Stick’s performance), we used generalized weighted quantile sum (gWQS) regression implemented in R (v4.5.2) using the gWQS package^51–53^. WQS is a mixtures-based ensemble modeling strategy that evaluates the joint effect of correlated exposures on an outcome of interest, while identifying each component’s relative contribution. The Cu mixture included concentrations measured in blood, urine, hair, and saliva. Pearson’s correlations between all Cu biomarkers collected in PHIME2-MRI were calculated to assess interrelationships among matrices. Before model estimation, all biomarkers were transformed into deciles.

The WQS analysis is implemented in two steps. First, a weighted index representing the association between each individual metal and the outcome is estimated across 50 bootstrap samples. Second, this weighted index is tested in a linear regression model predicting the association between the mixture and the outcome. These two steps are repeated multiple times to obtain more robust results. In the present study, we performed 100 repeated holdouts. The mixture is defined such that:

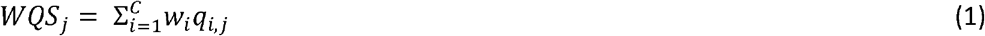

where *w*_*i*_ represents the empirically estimated weight for each predictor variable (i) and *q*_*i*_ denotes the ranked concentration of that predictor per subject (j). A significance test for the WQS index provides an estimate of the association with the overall mixture, while the weights associated with each predictor provide an indicator of each individual variable’s contribution to the overall effect. All weights are constrained to sum to one, enabling sorting by relative importance. Biological matrices that impact the outcome have larger weights; factors with little or no impact on the outcome have near-zero weights. These models were adjusted for age and sex.

Because active and passive smoking have been shown to affect OB structure and olfactory performance^54,55^, smoking exposure was accounted for in sensitivity analyses. Self-reported smoking information was collected through a study-specific Smoking, Alcohol, Coffee and Tea questionnaire which assessed active smoking and passive exposure within the home. We created a composite variable reflecting overall smoking exposure. Smoking data were available for 167 of 200 participants; after integrating biomarker, MRI, demographic, and smoking information, 161 participants had complete data for the sensitivity analyses.

## 4. Results

### 4.1. Demographic and exposure characteristics

This study included 200 participants (64% female, mean age 19.11 years (SD = 2.44)) living in the province of Brescia in Northern Italy. Cu concentrations in all four media are reported in Table 1. Correlations between Cu concentrations in each biological matrix ranged from −0.11 to 0.03 (Pearson’s r, p < 0.050; Supplementary Material Figure S2).

### 4.2. Cu exposure and olfactory bulb volume

Using WQS analyses, we observed significant negative association between the Cu exposure index and all OB volumetric measures associations (β_LEFT_ = −0.72, 95% CI [−1.42, −0.02], β_RIGHT_= −0.79, 95% CI [−1.43, −0.15], β_TOTAL_ = −1.55, 95% CI [−2.85, −0.25]; Figure 1). Across all models, similar matrices emerged as the strongest contributors the Cu mixture: hair and blood contributed most to the left OB volume (33% and 29%, respectively), urine and hair contributed most to the right OB volume (32% and 31%, respectively), urine and hair contributed most to total OB volume (32% and 31%, respectively).

**Figure 1.**
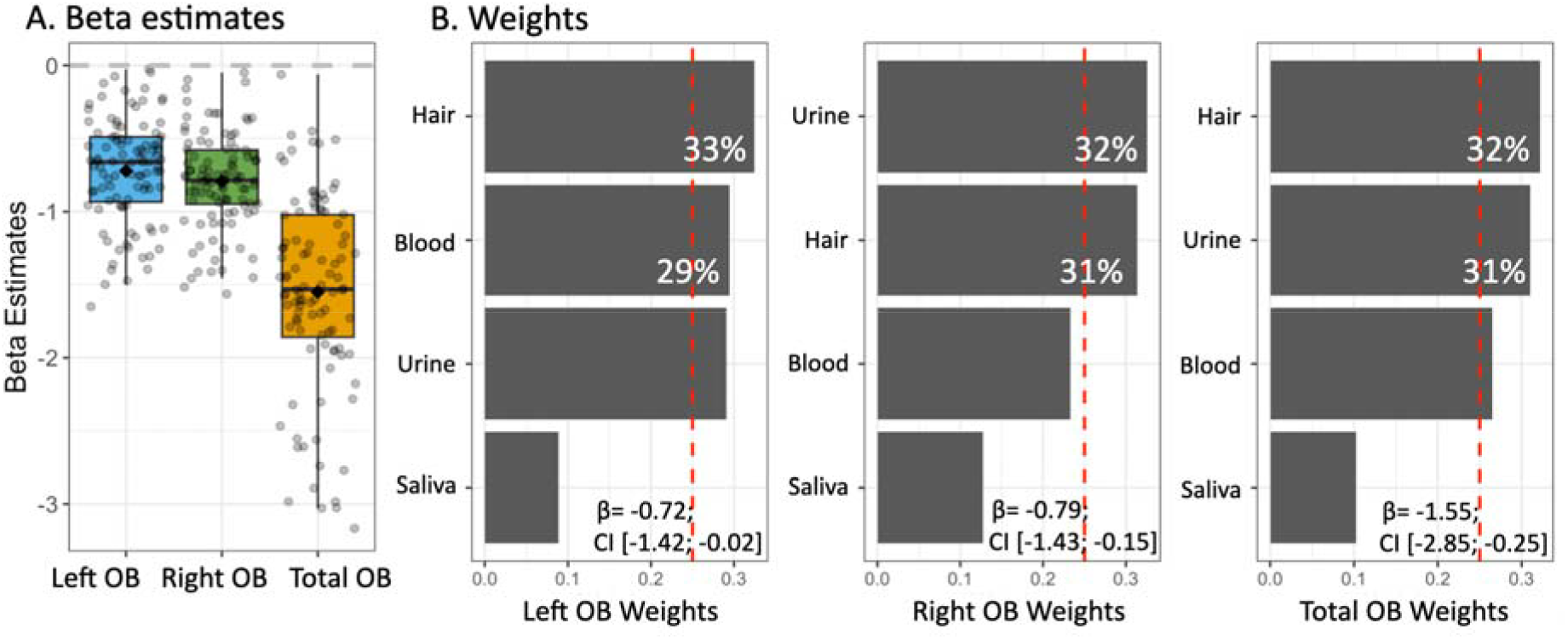
Copper exposure index associated with reduced olfactory bulb volume (n = 200). (A) Beta estimates from WQS models show significant negative associations between the Cu mixture index and left, right, and total OB volumes. Box plots display the distribution of beta estimates across 100 repeated holdouts (median and interquartile range), with whiskers showing the 10th and 90th percentiles. (B) Relative weights for each Cu biomarker (blood, urine, hair, saliva) derived from the WQS models illustrate each matrix’s contribution to the overall mixture effect. Larger weights indicate a greater influence of that biomarker on the outcome. The red dotted line represents the threshold that corresponds to the scenario where all elements contribute equally to the association. Models were adjusted for sex and age.

Sensitivity analyses adjusting for smoking yielded no significant associations (Supplementary Figure S3). In addition, standard linear regression models using individual Cu biomarkers did not identify significant associations with OB volumes (Supplementary Material Table S3; Figure S4).

### 4.3. Cu exposure and olfactory performance

We examined associations between the Cu exposure index and Sniffin’ Sticks outcomes using WQS. A significant negative association was found between the Cu exposure index and the threshold subtest (β_THRESHOLD_ = −0.23, 95% CI [−0.42, −0.03]; Figure 2). Blood and saliva were the primary contributors to this mixture effect, accounting for 42% and 40% of the overall weight, respectively (Figure 2). We did not observe significant associations between the Cu exposure index and other Sniffin’ Sticks outcomes (Discrimination, Identification, and TDI scores) (Supplementary Figure S5). For completeness, we evaluated linear associations between each individual biological matrix with each Sniffin’ Sticks outcome; none remained significant after multiple comparisons corrections (Supplementary Table S4 and Figure S4). Sensitivity analyses adjusting for smoking yielded no significant results (Supplementary Figure S6).

**Figure 2.**
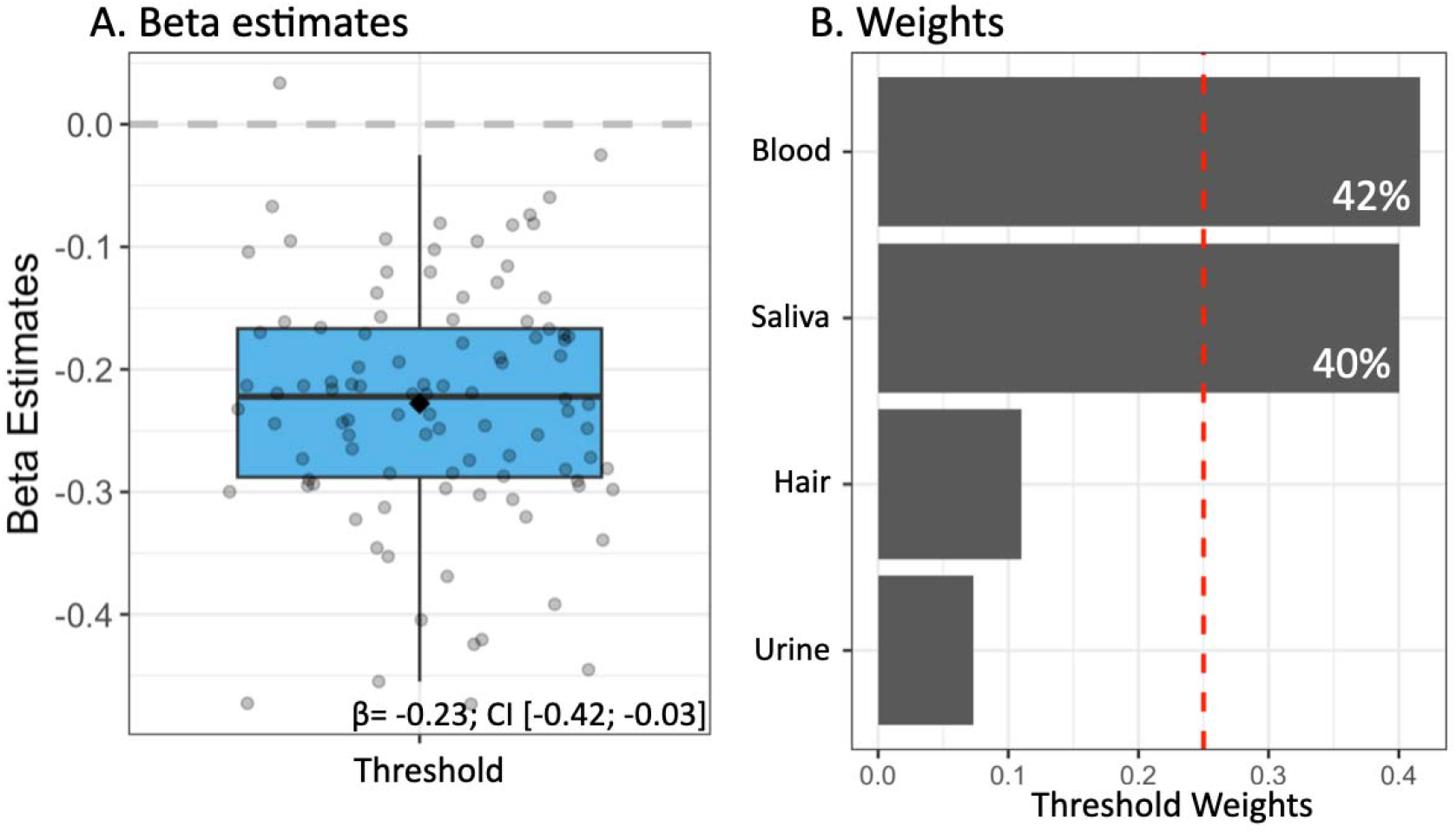
Copper exposure index associated with reduced Sniffin’ Sticks threshold subtest scores (n = 200). (A) Beta estimates from the WQS model show significant negative associations between the Cu mixture index and threshold subtest scores. Box plots display the distribution of beta estimates across 100 repeated holdouts (median and interquartile range), with whiskers showing the 10th and 90th percentiles. (B) Relative weights for each Cu biomarker (blood, urine, hair, and saliva) derived from the WQS models illustrate each matrix’s contribution to the overall mixture effect. Larger weights indicate a greater influence of that biomarker on the outcome. The red dotted line represents the threshold that corresponds to the scenario where all elements contribute equally to the association. Models were adjusted for sex and age.

## 5. Discussion

In this study, we provide the first epidemiologic evidence that multi-matric Cu exposure is associated with olfactory bulb structure and olfactory performance during adolescence. By integrating deep learning derived olfactory bulb volumes, quantitative olfactory testing, and a mixture-based exposure framework, our findings identify the olfactory system as a sensitive developmental target of environmental Cu exposure. A higher Cu index was consistently associated with reduced OB volume across the left, right, and total measures, and negatively associated with olfactory threshold performance, a behavioral measure linked to OB processing. For both structural and behavioral outcomes, hair, blood, saliva contributed differentially to the overall mixture effect, underscoring the value of a multi-matrix assessment of Cu exposure. Notably, individual Cu biomarkers were not associated with OB structure or olfactory performance when examined separately, highlighting the importance of considering cumulative Cu exposure. Together, these findings provide novel evidence that adolescent Cu exposure may adversely influence OB structure and olfactory performance during a sensitive window of development.

The relative weights in our WQS models suggest that both long-term Cu exposure, captured by hair Cu, and the body’s excretory response to ongoing exposure, reflected by urinary Cu, contribute to OB structural changes in adolescence. Hair Cu is widely used as an indicator of chronic exposure, reflecting metal accumulation over months,^44,56,57^ whereas urinary Cu primarily reflects more recent systemic exposures and excretion^58^. Structural changes to the brain are likely to occur gradually, suggesting that these associations are attributed to chronic Cu exposure rather than short-term changes in exposure. Although some lateral volumetric differences were observed with blood Cu emerging as a leading contributor in the left OB volume association with the Cu exposure index, hair remained the dominant contributor, reinforcing the likelihood that longer-term Cu dysregulation affects the olfactory system anatomy.

In contrast, salivary and blood Cu contributed most to associations with olfactory performance, specifically odor threshold. Because blood and saliva Cu levels are tightly regulated by homeostatic absorption, transport, and excretion mechanisms, they are imperfect biomarkers for short term exposure fluctuations^59,60^. Nonetheless, subtle increases in Cu concentrations in these matrices may occur when homeostatic systems are strained^59,60^. Even minor disruptions in Cu metabolism can affect neuronal activity and synaptic transmission, as Cu plays a key role in mitochondrial function and neurotransmitter regulation^61,62^. This pattern is consistent with our findings: elevated Cu mixture levels were associated with reduced olfactory threshold sensitivity, suggesting early impairment in the detection and transmission of odor information along the olfactory pathway. Given that different biological matrices reflect distinct exposure windows, these findings may indicate that shorter term variations in Cu levels (captured by blood and saliva) may be related to transient changes in olfactory performance, particularly odor threshold, whereas biomarkers reflecting longer term exposure (e.g., hair, urine) are more strongly related to structural differences in the OB. However, these interpretations require confirmation in longitudinal studies. Taken together, these results suggest that Cu neurotoxicity in the olfactory system may involve both differences in OB structure and alterations in olfactory sensory performance across exposure timescales.

Our observation of Cu-related reductions in OB volume and olfactory performance align with experimental evidence from animal models showing that Cu exposure impairs odor detection, disrupts olfactory-dependent behaviors, and alters sensory processing in fish and rodents^21–23^. Similarly, human neuroimaging studies demonstrate that reduced OB volume is associated with impaired olfaction and is a key early feature of neurodegenerative diseases such as Alzheimer’s and Parkinson’s disease^63,64^. The threshold subtest of Sniffn Sticks reflects odor detection sensitivity (e.g., how low a concentration of an odor one can detect); lower scores indicate lower olfactory sensitivity (e.g., a higher concentration of the odorant was needed to detect it). Our findings show that increased Cu exposure was linked to lower olfactory sensitivity, suggesting early impairment in the ability to detect and process odors. These findings are supported by studies that have shown with increasing age, Sniffin’ Sticks threshold subtest scores decline more steeply than the identification and discrimination subtest scores^12,65^. Impaired peripheral olfactory sensitivity is one of the earliest clinical indicators of neurodegenerative diseases, particularly Parkinson’s disease, where olfactory loss precedes motor symptoms by years^66^, our findings raise concern that Cu exposure during adolescence may set the stage for subtle but meaningful alterations in olfactory system integrity.

To contextualize Cu levels in our cohort, it is important to note that there are no formally established reference values for Cu concentration in biological matrices in Italy, and available values vary by region, lifestyle, age, and sex. In Italian adults, reported reference ranges for blood Cu range from 776–1495 μg/L (GM 1036 μg/L) in Sardinia^67^ to 807–1643 μg/L (mean 1225 μg/L) in Northern Italy^68^. A pediatric study in the United States established reference values of 570–1290 μg/L for those > 12.5 years, GM = 1030 ug/L for 0.5 - 18 years^69^. Our adolescent cohort’s blood Cu concentration GM (586 ng/mL = 586 μg/L) is lower than reference values reported in the literature. However, cross-study comparisons should be interpreted cautiously given differences in age distribution, environmental sources, and differences in Cu metabolism across developmental stages.

## 6. Limitations

A strength of this cross-sectional study is the use of a novel, well-characterized dataset that integrates neuroimaging, quantitative olfactory assessment, and multi-matrix copper biomarkers, enabling the simultaneous examination of brain structure and sensory function in adolescents. While we observed significant associations between the Cu mixture and both brain and behavior outcomes, our modest sample size limits statistical power. Replication in larger cohorts would strengthen confidence in our findings. However, to our knowledge, datasets in this age group that combine high resolution MRI scans, multiple biological markers, and detailed behavioral phenotypes are limited in availability, underscoring the unique value of this cohort. We also looked at a single metal and we are aware of other metals that are olfactory neurotoxicants. Given the relatively low Cu levels observed in our population, it is possible that the detected associations reflect unmeasured correlated exposures rather than copper alone. Additionally, we measured total blood Cu rather than ceruloplasmin-free Cu. Because most circulating Cu is bound to ceruloplasmin, a copper-transport protein involved in Cu homeostasis, total blood Cu may not fully capture the unbound Cu that can cross the blood brain barrier and contribute to neurotoxicity as observed in Wilson’s Disease^70–72^. Further investigations should evaluate multi-metal exposure profiles and metal mixtures to better disentangle causal agents and characterize the combined neurodevelopmental impacts of environmental metal exposures.

## 7. Conclusion

By applying our WQS approach, we identified associations between a multi-matrix Cu exposure index and both OB structure and olfactory performance in adolescents, with effects on olfactory performance driven primarily by olfactory sensitivity. Integration of Cu biomarkers across multiple biological matrices enabled capture of complementary information reflecting distinct exposure timeframes and physiological processes. Our findings that hair and urinary Cu contributed most to OB volume whereas saliva and blood Cu contributed most to the association with olfactory sensitivity and Cu exposure index, highlight the value of multi-matrix biomarker assessment in capturing gradual structural changes in the brain and physiological changes related to olfactory performance. Linear models assessing individual biomarkers and the Cu index were not significant, suggesting that changes in OB volume and olfactory outcomes may reflect the combined influence of multiple exposure pathways rather than any single biological matrix. Our results suggest that alterations in OB structure and olfactory performance during adolescence may represent early indicators of neural vulnerability. Few studies have examined early biomarkers during this critical development window, and our findings highlight the importance of investigating environmental exposures that may influence neural pathways relevant to long term neurological health, including neurodegenerative risk.

## Data Availability

All data produced in the present study are available upon reasonable request to the authors

## Data Availability

De-identified data will be made available upon reasonable request to the corresponding author. Defaced image files can be accessed upon completion of a data use agreement.

## Acknowledgements

The authors would like to acknowledge support from the National Institute of Environmental Health Sciences (NIEHS) grants numbers R01 ES019222, R01 ES028927, R01 ES013744, P30ES023515, and the European Union through its Sixth Framework Programme for RTD (contract number FOOD-CT-2006-016253).

## APPENDIX A Supplementary data

